# Comparing 3D, 2.5D, and 2D Approaches to Brain Image Segmentation

**DOI:** 10.1101/2022.11.03.22281923

**Authors:** Arman Avesta, Sajid Hossain, MingDe Lin, Mariam Aboian, Harlan M. Krumholz, Sanjay Aneja

## Abstract

Deep-learning methods for auto-segmenting brain images either segment one slice of the image (2D), five consecutive slices of the image (2.5D), or an entire volume of the image (3D). Whether one approach is superior for auto-segmenting brain images is not known.

We compared these three approaches (3D, 2.5D, and 2D) across three auto-segmentation models (capsule networks, UNets, and nnUNets) to segment brain structures. We used 3430 brain MRIs, acquired in a multi-institutional study, to train and test our models. We used the following performance metrics: segmentation accuracy, performance with limited training data, required computational memory, and computational speed during training and deployment.

3D, 2.5D, and 2D approaches respectively gave the highest to lowest Dice scores across all models. 3D models maintained higher Dice scores when the training set size was decreased from 3199 MRIs down to 60 MRIs. 3D models converged 20% to 40% faster during training and were 30% to 50% faster during deployment. However, 3D models require 20 times more computational memory compared to 2.5D or 2D models.

This study showed that 3D models are more accurate, maintain better performance with limited training data, and are faster to train and deploy. However, 3D models require more computational memory compared to 2.5D or 2D models.

## Introduction

Segmentation of brain magnetic resonance images (MRIs) has widespread applications in the management of neurological disorders.^1–3^ In patients with neurodegenerative disorders, segmenting brain structures such as the hippocampus provides quantitative information about the amount of brain atrophy.^4^ In patients undergoing radiotherapy, segmentation is used to demarcate important brain structures that should be avoided to limit potential radiation toxicity.^5^ Pre-operative or intra-operative brain MRIs are often used to identify important brain structures that should be avoided during neurosurgery.^6,7^ Manual segmentation of brain structures on these MR images is a time-consuming task that is prone to intra- and inter-observer variability.^8^ As a result, deep learning auto-segmentation methods have been increasingly used to efficiently segment important anatomical structures on brain MRIs.^9^

Compared to two-dimensional (2D) auto-segmentation tasks, the three-dimensional (3D) nature of brain MRIs makes auto-segmentation considerably more challenging (Figure 1). There have been three proposed approaches to handling auto-segmentation of 3D images: 1) analyze and segment a two-dimensional slice of the image at a time (2D),^10^ 2) analyze five consecutive two-dimensional slices at a time to generate a segmentation of the middle slice (2.5D),^11^ and 3) analyze and segment the image volume in the three-dimensional space (3D).^10^ Although each approach has shown some promise in medical image segmentation, a comprehensive comparison and benchmarking of these approaches for auto-segmentation of brain MRIs is lacking. Prior studies on comparing these auto-segmentation approaches have often not evaluated their efficacy in segmenting brain MRIs, or have limited their comparison narrowly to one deep learning architecture. ^10,12–14^ Additionally, previous studies have focused primarily on segmentation accuracy and failed to evaluate more practical metrics such as computational efficiency or accuracy in data-limited settings. As a result, it is difficult for clinicians and researchers to easily choose the appropriate auto-segmentation method for a desired clinical task. There is a need to compare and benchmark these three approaches for brain MRI auto-segmentation across different models and using comprehensive performance metrics.

**Figure 1:**
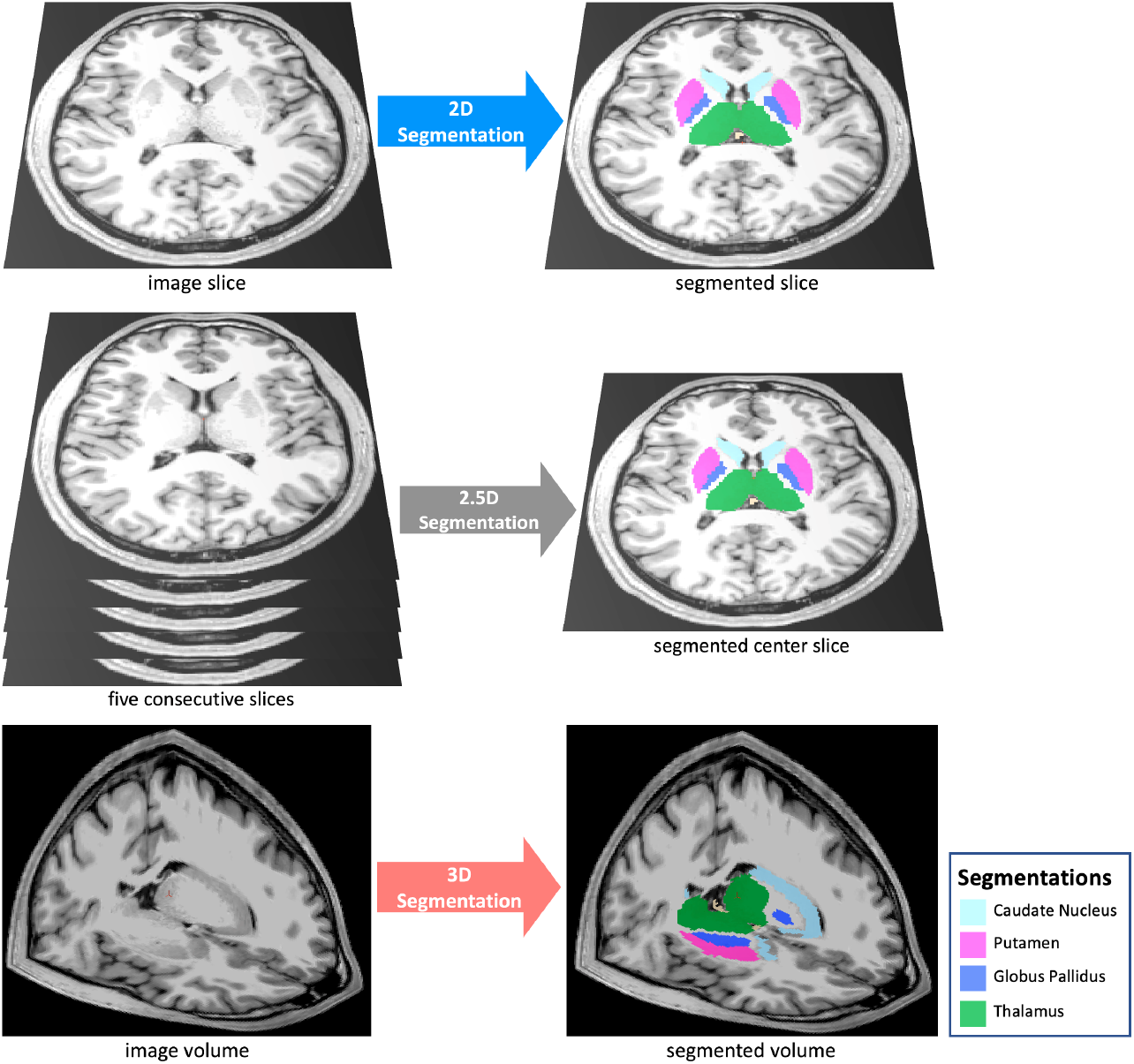
we compared three segmentation approaches (3D, 2.5D, and 2D) across three auto-segmentation models (capsule networks, UNets, and nnUNets). The 2D approach analyzes and segments one slice of the image, the 2.5D approach analyzes five consecutive slices of the image to segment the middle slice, and the 3D approach analyzes and segments a 3D volume of the image.

In this study, we comprehensively compared 3D, 2.5D, and 2D approaches to brain MRI auto-segmentation across three different deep learning architectures and used metrics of accuracy and computational efficiency. We used a multi-institutional cohort of 3,430 brain MRIs to train and test our models, and evaluated the efficacy of each approach across three clinically-relevant anatomical structures of the brain.

## Methods

### Dataset

This study used a dataset of 3,430 T1-weighted brain MR images belonging to 841 patients from 19 institutions enrolled in the Alzheimer’s Disease Neuroimaging Initiative (ADNI) study.^15^ The inclusion and exclusion criteria of ADNI have been previously described.^16^ On average, each patient underwent four MRI acquisitions. Each patient underwent MR imaging using a single scanner at each site. However, the diversity of scanners in all study sites included nine different types of MR scanners. Appendix 1 describes the details of MRI acquisition parameters. We downloaded the anonymized MRIs of these patients from Image and Data Archive, which is a data-sharing platform.^15^ The patients were randomly split into training (3,199 MRIs, 93% of data), validation (117 MRIs, 3.5% of data), and test (114 MRIs, 3.5% of data) sets *at the patient level*. Therefore, all images belonging to a patient were assigned to either the training, validation, or test set. Table 1 summarizes patient demographics.

**Table 1:**
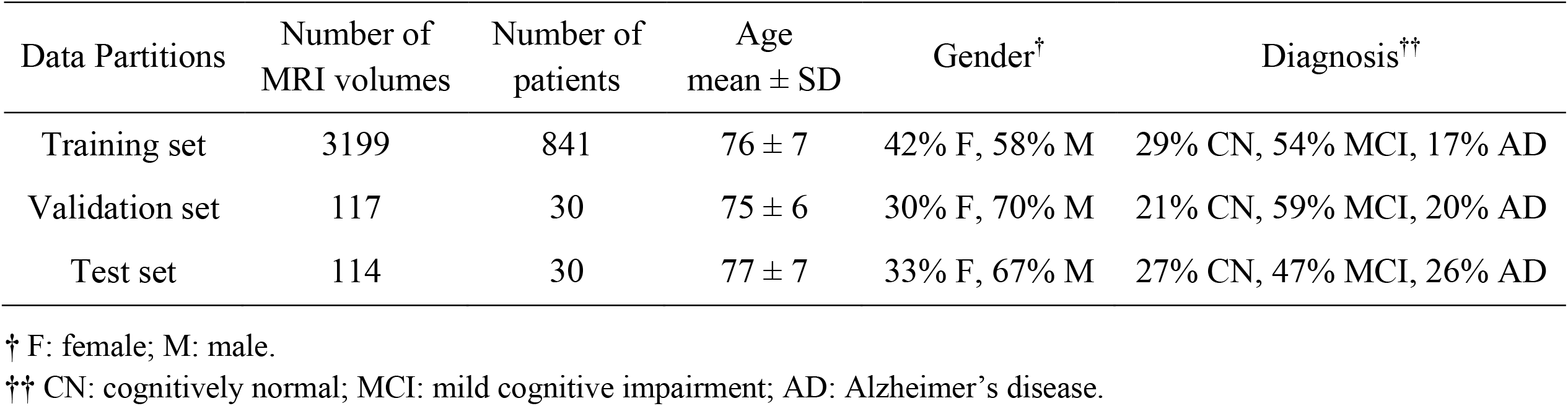
Study participants. tabulated by the training, validation, and test sets.

| Data Partitions | Number of<br>MRI volumes | Number of<br>patients | Age<br>mean ± SD | Gender<sup>†</sup> | Diagnosis<sup>††</sup> |
| --- | --- | --- | --- | --- | --- |
| Training set | 3199 | 841 | 76 ± 7 | 42% F, 58% M | 29% CN, 54% MCI, 17% AD |
| Validation set | 117 | 30 | 75 ± 6 | 30% F, 70% M | 21% CN, 59% MCI, 20% AD |
| Test set | 114 | 30 | 77 ± 7 | 33% F, 67% M | 27% CN, 47% MCI, 26% AD |

<sup>†</sup> F: female; M: male.
<sup>††</sup> CN: cognitively normal; MCI: mild cognitive impairment; AD: Alzheimer’s disease.

### Anatomic Segmentations

We trained our models to segment three representative structures of the brain: the third ventricle, thalamus, and hippocampus. These structures represent varying degrees of segmentation difficulty: the third ventricle is an easy structure to segment because it is filled with cerebrospinal fluid (CSF) with a distinct image contrast compared to surrounding structures; the thalamus is a medium-difficulty structure because it is bounded by CSF on one side and is bounded by white-matter on the other side, and the hippocampus is a difficult structure because it has a complex shape and is neighbored by multiple brain structures with different image contrasts. Preliminary ground-truth segmentations were initially generated by FreeSurfer,^4,17,18^ and were manually corrected by a board-eligible radiologist (AA).

### Image Pre-Processing

MRI preprocessing included corrections for B1-field variations as well as intensity inhomogeneities.^19,20^ The 3D brain image was cropped around the brain after removing the skull, face, and neck tissues.^21^ The input to the 3D capsule networks and 3D UNets were image patches sized at 64×64×64 voxels. The inputs to the 2.5D capsule networks and 2.5D UNets were five consecutive slices of the image. The inputs to the 2D capsule networks and 2D UNets were one slice of the image. The inputs to the 3D and 2D nnUNet models were respectively 3D and 2D patches of the images with self-configured patch sizes that were automatically set by the nnUNet paradigm.^22^ Appendix 2 describes the details of pre-processing.

### Auto-segmentation Models

We compared the 3D, 2.5D, and 2D approaches across three segmentation models: capsule networks (CapsNets),^23^ UNets,^24^ and nnUNets.^22^ These models are considered the highest-performing auto-segmentation models in the biomedical domain.^9,22,23,25–29^ The 3D models process a 3D patch of the image as input, all feature maps and parameter tensors in all layers are 3D, and the model output is the segmented 3D patch of the image. Conversely, 2D models process a 2D slice of the image as input, all feature maps and parameter tensors in all layers are 2D, and the model output is the segmented 2D slice of the image. The 2.5D models process five consecutive slices of the image as input channels. The rest of the 2.5D model, including the feature maps and parameter tensors, are 2D, and the model output is the segmented 2D middle slice among the five slices. We did not develop 2.5D nnUNets, because the self-configuring paradigm of nnUNets is developed for 3D and 2D inputs but not for 2.5D inputs. Notably, the aim of training and testing nnUNets (in addition to UNets) was to ensure that our choices of hyperparameters did not cause one approach (such as 3D) to perform better than other approaches. The nnUNet can self-configure the best hyperparameters for 3D and 2D approaches but not for the 2.5D approach. As a result, we did not train or test 2.5D nnUNets. The model architectures are described in Appendix 3.

### Training

We trained the CapsNet and UNet models for 50 epochs using Dice loss and the Adam optimizer.^30^ Initial learning rate was set at 0.002. We used dynamic paradigms for learning rate scheduling, with a minimal learning rate of 0.0001. The hyperparameters for our CapsNet and UNet models were chosen based on the model with the lowest Dice loss over the validation set. The hyperparameters for the nnUNet model were self-configured by the model.^22^ Appendix 4 describes the training hyperparameters for CapsNet and UNet.

### Performance Metrics

For each model (CapsNet, UNet, and nnUNet), we compared the performance of 3D, 2.5D, and 2D approaches using the following metrics: 1) Segmentation accuracy: we used the Dice score to quantify the segmentation accuracy of the fully trained models over the test set.^31^ We compared Dice scores between the three approaches for three representative anatomic structures of the brain: the third ventricle, thalamus, and hippocampus. The mean Dice scores for the auto-segmentation of these brain structures are reported together with their 95% confidence interval. To compute the 95% confidence interval for each Dice score, we used bootstrapping to sample the 114 Dice scores over the test set, with replacement, 1000 times. We then calculated the mean Dice score for each of the 1000 samples, giving us 1000 mean Dice scores. We then sorted these mean Dice scores and found the range that covered 95% of them, which is equivalent to the 95% confidence interval for each Dice score; 2) Performance when training data is limited: we trained the models using the complete training set and random subsets of the training set with 600, 240, 120, and 60 MR images. The models trained on these subsets were then evaluated over the test set; 3) Computational speed during training: we compared the time needed to train the 3D, 2.5D, and 2D models per training example per epoch until the model converged; 4) Computational speed for segmenting an MR image: we compared how quickly each of the 3D, 2.5D, and 2D models segment one brain MRI volume; and 5) Computational memory: we compared how much GPU memory is required, in units of megabytes, to train and deploy each of the 3D, 2.5D, and 2D models.

### Implementation

Image pre-processing was done using Python (version 3.10) and FreeSurfer (version 7). PyTorch (version 1.12) was used for model development and testing. Training and testing of the models were run on GPU-equipped servers (4 vCPUs, 16 GB RAM, 16 GB NVIDIA GPU). The code used to train and test our models is available on our lab’s GitHub page: https://github.com/Aneja-Lab-Yale/Aneja-Lab-Public-3D2D-Segmentation.

## Results

The segmentation accuracy of the 3D approach across all models and all anatomic structures of the brain was higher than 2.5D or 2D approaches, with Dice scores of the 3D models above 90% for all anatomic structures (Table 2). Within the 3D approach, all models (CapsNet, UNet, and nnUNet) performed similarly in segmenting each anatomic structure, with their Dice scores within 1% of each other. For instance, the Dice scores of 3D CapsNet, UNet, and nnUNet in segmenting the hippocampus were respectively 92%, 93%, and 93%. Figure 2 shows auto-segmented brain structures in one patient using the three approaches.

**Table 2:** Comparing the segmentation accuracy of 3D, 2.5D, and 2D approaches across three auto-segmentation models to segment brain structures. The three auto-segmentation models included CapsNet, UNet, and nnUNet. These models were used to segment three representative brain structures: 3rd ventricle, thalamus, and hippocampus, which respectively represent easy, medium, and difficult structures to segment. The segmentation accuracy was quantified by Dice scores over the test (114 brain MRIs).

| CapsNet |  |  |  |
| --- | --- | --- | --- |
| Brain structure | 3D Dice (95% CI) | 2.5D Dice (95% CI) | 2D Dice (95% CI) |
| 3rd ventricle | 95 %<br>(94 to 96) | 90 %<br>(89 to 91) | 90 %<br>(88 to 92) |
| Thalamus | 94%<br>(93 to 95) | 76 %<br>(74 to 78) | 75 %<br>(72 to 78) |
| Hippocampus | 92 %<br>(91 to 93) | 73 %<br>(71 to 75) | 71 %<br>(68 to 74) |

| UNet |  |  |  |
| --- | --- | --- | --- |
| Brain structure | 3D Dice (95% CI) | 2.5D Dice (95% CI) | 2D Dice (95% CI) |
| 3rd ventricle | 96 %<br>(95 to 97) | 92%<br>(91 to 93) | 91 %<br>(89 to 91) |
| Thalamus | 95 %<br>(94 to 96) | 92%<br>(91 to 93) | 90 %<br>(88 to 92) |
| Hippocampus | 93 %<br>(92 to 94) | 86 %<br>(84 to 88) | 88 %<br>(86 to 90) |

| nnUNet |  |  |  |
| --- | --- | --- | --- |
| Brain structure | 3D Dice (95% CI) | 2.5D Dice (95% CI) | 2D Dice (95% CI) |
| 3rd ventricle | 96 %<br>(95 to 97) | N/A | 90 %<br>(89 to 91) |
| Thalamus | 94 %<br>(93 to 95) | N/A | 92 %<br>(91 to 93) |
| Hippocampus | 93 %<br>(92 to 94) | N/A | 91 %<br>(90 to 92) |

**Figure 2:**
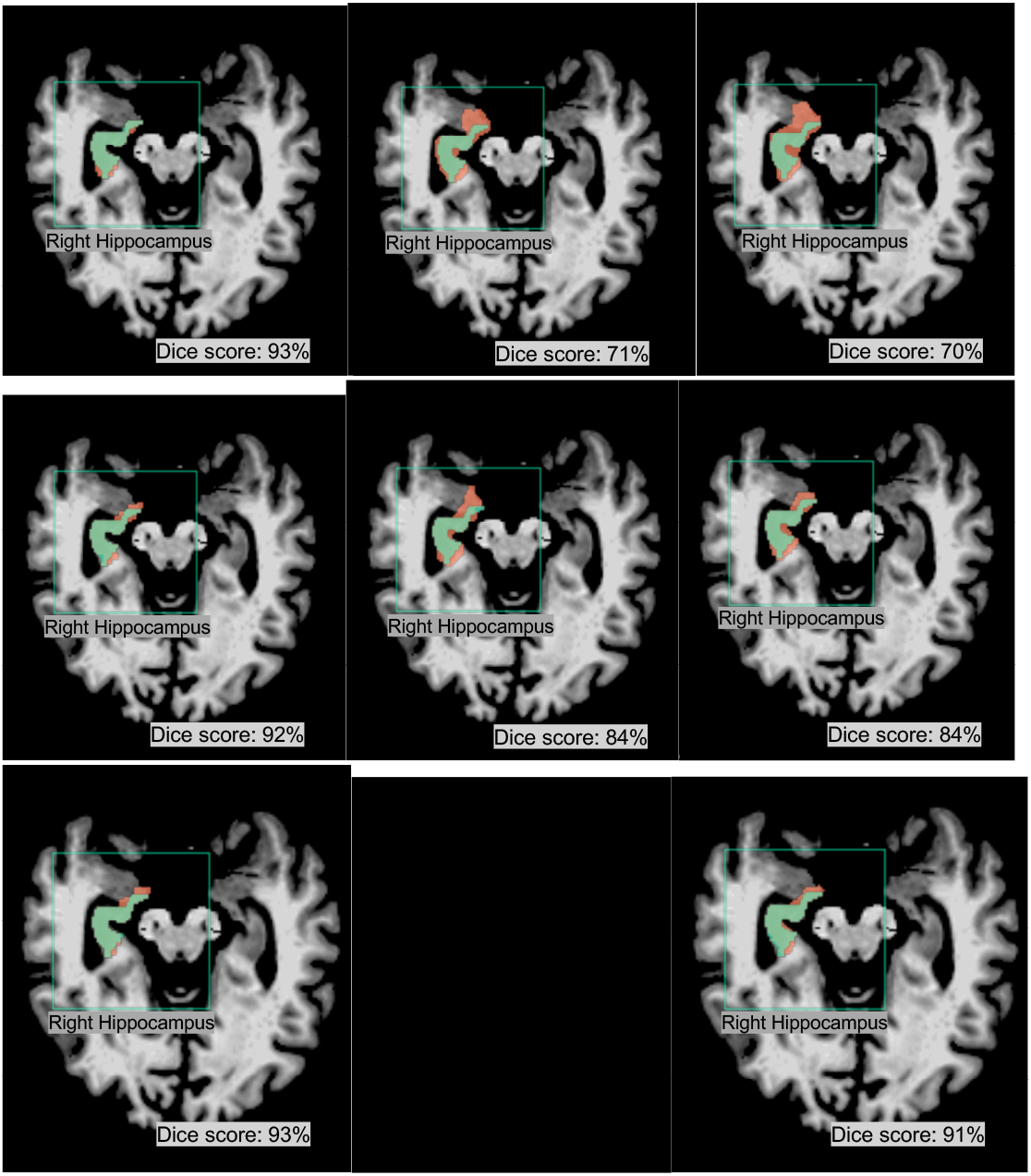
Examples of 3D, 2.5D, and 2D segmentations of the right hippocampus by CapsNet, UNet, and nnUNet. Target segmentations and model predictions are respectively shown in green and red. Dice scores are provided for the entire *volume* of the right hippocampus *in this patient* (who was randomly chosen from the test set).

3D models maintained higher accuracy, compared to 2.5D and 2D models, when training data was limited (Figure 3). When we trained the 3D, 2.5D, and 2D CapsNets using the full training set containing 3,199 MRIs, their Dice scores in segmenting the third ventricle were respectively 95%, 90%, and 90%. When we trained the same models on smaller subsets of the training set containing 600, 240, 120, and 60 MRIs, the performance of 3D, 2.5D, and 2D CapsNets gradually decreased down to 90%, 88%, and 87% for the 3D, 2.5D, and 2D CapsNets, respectively (Figure 3). Importantly, the 3D CapsNet maintained higher Dice scores (over the test set) compared to 2.5D or 2D CapsNets in all these experiments. Similarly, when we trained 3D, 2.5D, and 2D UNets using the full training set, their Dice scores in segmenting the third ventricle were respectively 96%, 91%, and 90%. Decreasing the size of the training set down to 60 MRIs resulted in Dice scores of 90%, 88%, and 87% for the 3D, 2.5D, and 2D UNets, respectively. Again, the 3D UNet maintained higher Dice scores compared to 2.5D or 2D UNets in all these experiments. Lastly, when we trained 3D and 2D nnUNets using the full training set, their Dice scores in segmenting the third ventricle were respectively 96% and 90%. Decreasing the size of the training set down to 60 MRIs resulted in Dice scores of 92% and 87% for the 3D and 2D nnUNets, respectively. Once more, the 3D nnUNet maintained higher Dice scores compared to the 2D nnUNet in all these experiments (Figure 3).

**Figure 3:**
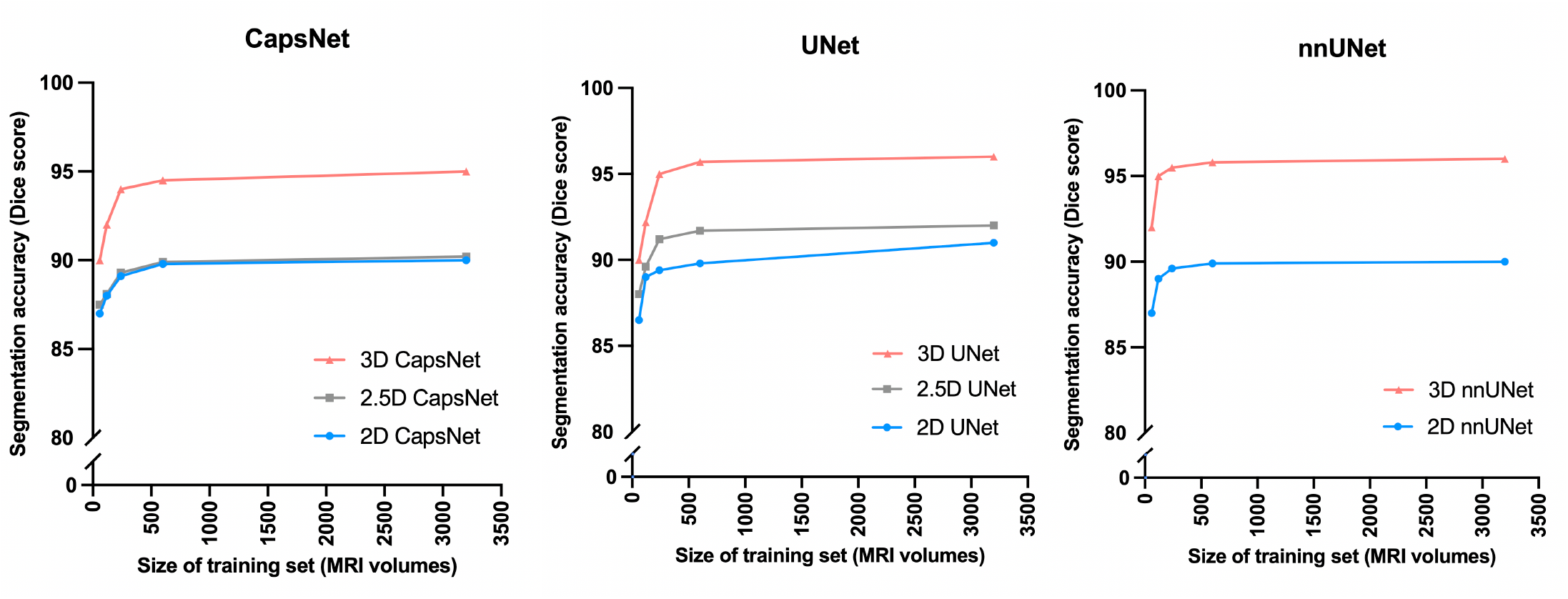
Comparing 3D, 2.5D, and 2D approaches when training data is limited. As we decreased the size of the training set from 3000 MRIs down to 60 MRIs, the 3D models maintained higher segmentation accuracy (measured by Dice scores).

The 3D models trained faster compared to 2.5D or 2D models (Figure 4). The 3D, 2.5D, and 2D CapsNets respectively took 0.8, 1, and 1 seconds per training example per epoch to converge during training. The 3D, 2.5D, and 2D UNets respectively took 1.6, 2.2 and 2.9 seconds per training example per epoch to converge during training. The 3D and 2D nnUNets respectively took 2 and 2.9 seconds per training example per epoch to converge during training. Therefore, 3D models converged 20% to 40% faster compared to 2.5D or 2D models.

**Figure 4:**
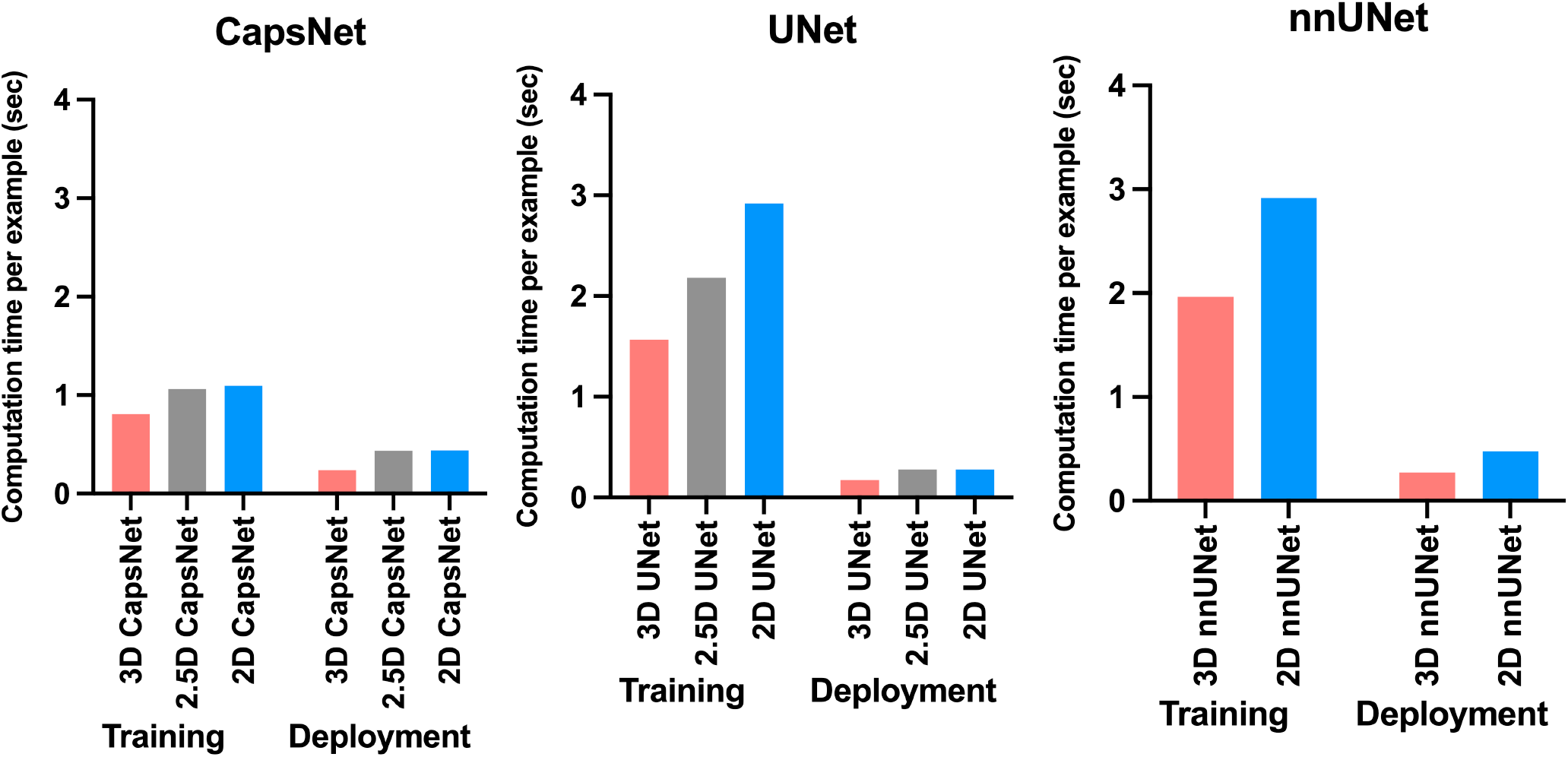
Comparing computational time required by 3D, 2.5D, and 2D approaches to train and deploy auto-segmentation models. The training times represent how much time it would take per training example per epoch for the model to converge. The deployment times represent how much time each model would require to segment one brain MRI volume. The 3D approach trained and deployed faster across all auto-segmentation models.

Fully-trained 3D models could segment brain MRIs faster during deployment compared to 2.5D or 2D models (Figure 4). Fully-trained 3D, 2.5D, and 2D CapsNets could respectively segment a brain MRI in 0.2, 0.4, and 0.4 seconds. Fully-trained 3D, 2.5D, and 2D UNets could respectively segment a brain MRI in 0.2, 0.3, and 0.3 seconds. Lastly, fully-grained 3D and 2D nnUNets could respectively segment a brain MRI in 0.3 and 0.5 seconds. Therefore, fully-trained 3D models segment a brain MRI 30% to 50% faster compared to fully-trained 2.5D or 2D models.

The 3D models need more computational memory to train and deploy as compared to 2.5D or 2D models (Figure 5). The 3D, 2.5D, and 2D CapsNets respectively require 317, 19, and 19 megabytes of GPU memory during training. The 3D, 2.5D, and 2D UNets respectively require 3150, 180, and 180 megabytes of GPU memory. The 3D and 2D nnUNets respectively require 3200 and 190 megabytes of GPU memory. Therefore, 3D models required about 20 times more GPU memory compared to 2.5D or 2D models. Notably, CapsNets require 10 times less GPU memory compared to UNets or nnUNets. Therefore, 3D CapsNets only required two times more GPU memory compared to 2.5D or 2D UNets or nnUNets (Figure 5).

**Figure 5:**
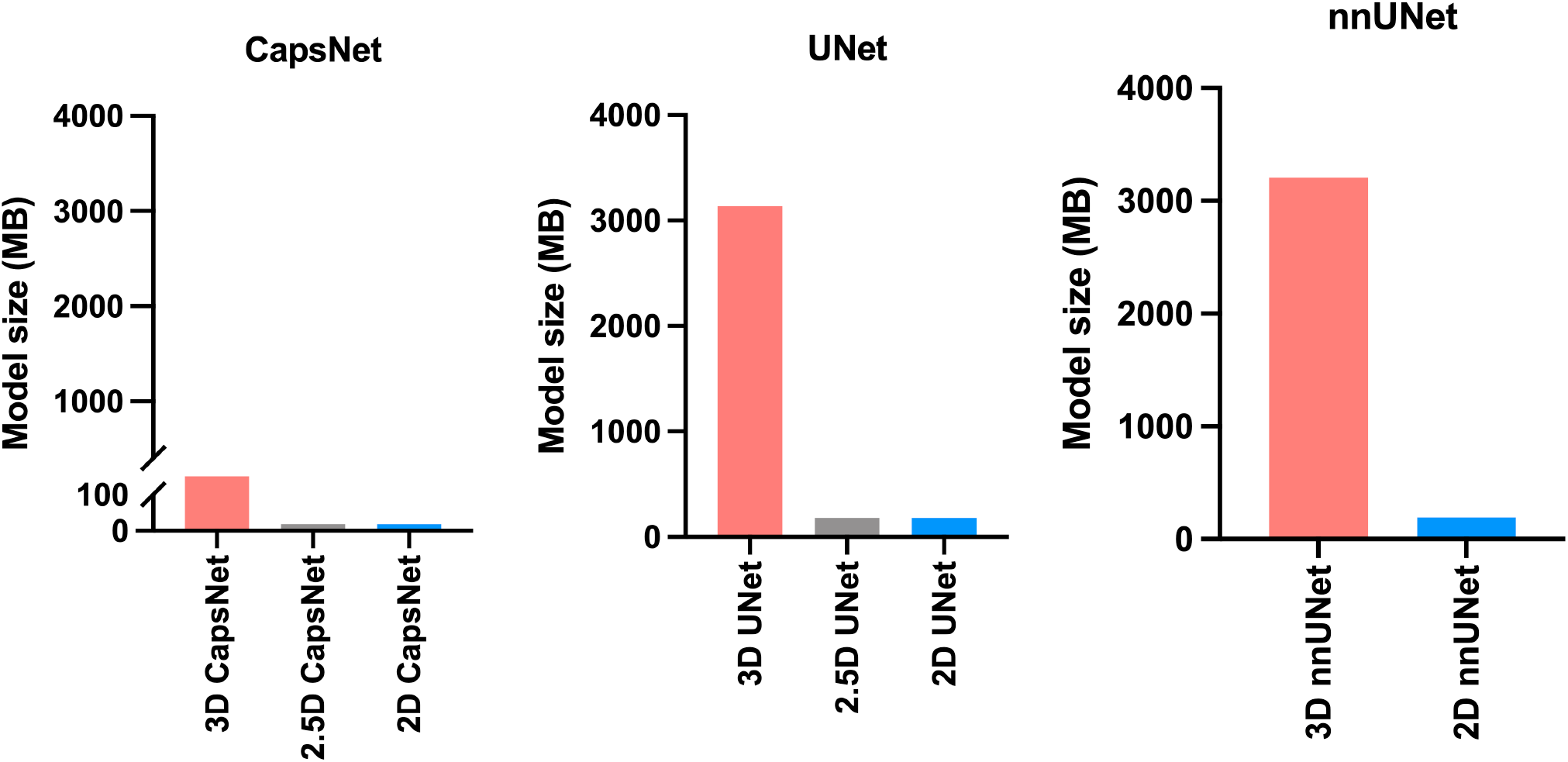
Comparing the memory required by 3D, 2.5D, and 2D approaches. The bars represent the computational memory required to accommodate the total size of each model, including the parameters plus the cumulative size of the forward- and backward-pass feature volumes. Within each auto-segmentation model (CapsNet, UNet, and nnUNet), the 3D approach requires 20 times more computational memory compared to 2.5D or 2D approaches.

## Discussion

In this study, we compared 3D, 2.5D, and 2D approaches of auto-segmentation across three different deep learning architectures, and found that the 3D approach is more accurate, faster to train, and faster to deploy. Moreover, the 3D auto-segmentation approach maintained better performance in the setting of limited training data. We found the major disadvantage of 3D auto-segmentation approaches to be increased computational memory requirement compared to similar 2.5D and 2D auto-segmentation approaches.

Our study extends the prior literature ^10,12,13,32,33^ in key ways. We provide the first comprehensive benchmarking of 3D, 2.5D, and 2D approaches in auto-segmenting brain MRIs, measuring both accuracy and computational efficiency. We compared 3D, 2.5D, and 2D approaches across three of the most successful auto-segmentation models to date, namely capsule networks, UNets, and nnUNets. Our findings provide a practical comparison of these three auto-segmentation approaches that can provide insight when attempting auto-segmentation in settings where computational resources are bounded or when the training data is limited.

We found that the 3D approache to auto-segmentation trains faster and deploys more quickly. Previous studies that compared the computational speed of 3D and 2D UNets have concluded conflicting results: some suggested that 2D models converge faster,^10,13,33^ while others suggested that 3D models converge faster.^22^ Notably, one training iteration of 2.5D or 2D models is faster than 3D models because 2.5D and 2D models have 20 times fewer trainable parameters compared to 3D models. However, feeding a 3D image volume into a 2.5D or 2D model requires a *for loop* that iterates through multiple slices of the image, which slows down 2.5D and 2D models. Additionally, 3D models can converge faster during training because they can use the contextual information in the 3D image volume to segment each structure.^10^ Conversely, 2.5D models can only use the contextual information in a few slices of the image,^11^ and 2D models can only use the contextual information in one slice only.^12^ Since the 3D approach provides more contextual information for each segmentation target, the complex shape of structures such as the hippocampus can be learned faster, and, as a result, the convergence of 3D models can become faster. Lastly, each training iteration through a 3D model can be accelerated by larger GPU memory, since the training of learnable parameters can be parallelized. However, each training iteration through a 2.5D or 2D model cannot be accelerated by larger GPU memory because iterations through the slices of the image (*for loop*) cannot be parallelized. We propose that our findings, that 3D models converge faster, are resulted from using state-of-the-art GPUs and efficient 3D models that learn contextual information in the 3D volume of the MR image faster. Our results also showed that the 3D models are faster during deployment since they can process the 3D volume of the image at once, while 2.5D or 2D models must loop through 2D image slices.

Our results did highlight one of the drawbacks of 3D auto-segmentation approaches. Specifically, we found that within each model, the 3D approach requires 20 times more computational memory compared to 2.5D or 2D approaches. Previous studies that compared 3D and 2D UNets have found similar results.^10,32^ This seems to be the only downside of the 3D approach compared to 2.5D or 2D approaches. Notably, the 2.5D approach was initially developed to achieve segmentation accuracy similar to the 3D approach while requiring computational resources similar to the 2D approach. In brain image segmentation, however, our results showed that the 2.5D approach could not achieve the segmentation accuracy of the 3D approach. This raises the question of which approach to use when computational memory is limited. Our results showed that *3D CapsNets* outperform all 2.5D and 2D models while only requiring twice more computational memory than the 2.5D or 2D UNets or nnUNets. Conversely, 3D UNets and nnUNets require 20 times more computational memory compared to 2.5D or 2D UNets and nnUNets. Therefore, 3D CapsNets may be preferred in settings where computational memory is limited.

Our results corroborate previous studies showing that deep learning is accurate in biomedical image auto-segmentation.^9,22,26–29^ Prior studies have shown that capsule networks, UNets, and nnUNets are the most accurate models to auto-segment biomedical images.^9,11,22,23,25,26,28,34,35^ Prior studies have also shown that the 3D, 2.5D, and 2D versions of these models can auto-segment medical images.^9,11,22,23,28,29^ However, evidence was lacking about which of the 3D, 2.5D, or 2D approaches would be more accurate in auto-segmenting brain structures on MR images. Our study also provides practical benchmarking of computational efficiency between the three approaches, which is often under-reported.

Our study has several notable limitations. First, we only compared the 3D, 2.5D, and 2D approaches to the auto-segmentation of brain structures on MR images. The results of this study may not generalize to other imaging modalities or other body organs. Second, there are multiple ways to develop a 2.5D auto-segmentation model.^11,36,37^ While we did not implement all the different versions of 2.5D models, we believe that our implementation of 2.5D models (using five consecutive image slices as input channels) is the best approach to segment the neuroanatomy on brain images. Third, our results about the relative deployment speed of 3D models as compared to 2.5D or 2D models might change as computational resources change. If the GPU memory is large enough to accommodate large 3D patches of the image, 3D models can segment the 3D volume faster. However, in settings where the GPU memory is limited, the 3D model should loop through multiple smaller 3D patches of the image, eroding the faster performance of the 3D models during deployment. However, we used a 16 GB GPU to train and deploy our models, which is commonplace in modern computing units used for deep learning. Finally, we compared 3D, 2.5D, and 2D approaches across three auto-segmentation models only: CapsNets, UNets, and nnUNets. While multiple other auto-segmentation models are available, we believe that our study has compared 3D, 2.5D, and 2D approaches across the most successful deep-learning models for medical image auto-segmentation. Further studies comparing the three approaches across other auto-segmentation models can be an area of future research.

## Conclusions

In this study, we compared 3D, 2.5D, and 2D approaches to brain image auto-segmentation across different models and concluded that the 3D approach is more accurate, achieves better performance in the context of limited training data, and is faster to train and deploy. Our results hold across various auto-segmentation models, including capsule networks, UNets, and nnUNets. The only downside of the 3D approach is that it requires 20 times more computational memory compared to the 2.5D or 2D approaches. Because 3D capsule networks only need twice the computational memory that 2.5D or 2D UNets and nnUNets need, we suggest using 3D capsule networks in settings where computational memory is limited.

## Data Availability

the data used in this study were obtained from the Alzheimer Disease Neuroimaging Initiative (ADNI) database available at: adni.loni.usc.edu.

https://adni.loni.usc.edu/

## Abbreviations

2D segmentation: two-dimensional segmentation
2.5D segmentation: enhanced two-dimensional segmentation
3D segmentation: three-dimensional segmentation
ADNI: Alzheimer’s disease neuroimaging initiative
CapsNet: capsule network
CPU: central processing unit
CT: computed tomography
GB: giga-byte
GPU: graphics processing unit
MRI: magnetic resonance imaging

## Author Contributions

conceptualization, methodology, validation, formal analysis, investigation, and visualization: Arman Avesta and Sanjay Aneja; software: Arman Avesta, Sajid Hossain, and Sanjay Aneja; resources: Arman Avesta, MingDe Lin, Mariam Aboian, Harlan Krumholz, and Sanjay Aneja; data curation: Arman Avesta and Mariam Aboian; writing—original draft preparation: Arman Avesta, Harlan Krumholz, and Sanjay Aneja; writing—review and editing: all co-authors; supervision: Mariam Aboina, Harlan Krumholz, and Sanjay Aneja; project administration: Arman Avesta and Sanjay Aneja; funding acquisition: Arman Avesta and Sanjay Aneja. All authors have read and agreed to the published version of the manuscript.

## Institutional Board Review Statement

this study was conducted in accordance with the Declaration of Helsinki, and approved by the Institutional Review Board of Yale School of Medicine (IRB number 2000027592, approved 4/20/2020).”

## Data Availability Statement

the data used in this study were obtained from the Alzheimer’s Disease Neuroimaging Initiative (ADNI) database (adni.loni.usc.edu). We obtained T1-weighted MRIs of 3,430 patients in the Alzheimer’s Disease Neuroimaging Initiative study from this data-sharing platform. The investigators within the ADNI contributed to the design and implementation of ADNI but did not participate in the analysis or writing of this article.

## Acknowledgments

None.

## Disclosures

**Arman Avesta:**

**Research Funding:** CTSA UL1 TR001863 from the National Center for Advancing Translational Science, and Fellow Research Grant Number RF2212 from the Radiological Society of North America.

**Potential Conflict of Interest:** Arman Avesta holds securities at Hyperfine Inc.

**Sajid Hossain:**

Nothing to disclose.

**MingDe Lin:**

MingDe Lin is an employee and stockholder of Visage Imaging, Inc. Unrelated to this work, MingDe is a board member of the Tau Beta Pi Engineering Honor Society.

**Mariam Aboian:**

**Research funding:** KL2 TR001862 from the National Center for Advancing Translational Science and NIH Roadmap for Medical Research. The contents of this article are solely the responsibility of the authors and do not necessarily represent the official view of NIH.

**Harlan M. Krumholz**

**Employment:** Hugo Health (I), FPrime

**Stock and Other Ownership Interests:** Element Science, Refactor Health, Hugo Health

**Consulting or Advisory Role:** UnitedHealthcare, Aetna

**Research Funding:** Johnson and Johnson

**Expert Testimony:** Siegfried and Jensen Law Firm, Arnold and Porter Law Firm, Martin/Baughman Law Firm

**Sanjay Aneja:**

**Research Funding:** The MedNet, Inc, American Cancer Society, National Science Foundation, Agency for Healthcare Research and Quality, National Cancer Institute, ASCO, The Patterson Trust

**Patents, Royalties, Other Intellectual Property:** Provisional patent of deep learning optimization algorithm

**Travel, Accommodations, Expenses:** Prophet Consulting (I), Hope Foundation

**Other Relationship:** NRG Oncology Digital Health Working Group, SWOG Digital Engagement Committee, ASCO mCODE Technical Review Group, Associate Editor for *JCO Clinical Cancer Informatics*

## SUPPLEMENTAL MATERIAL

## Appendix 1

MRI acquisition parameters

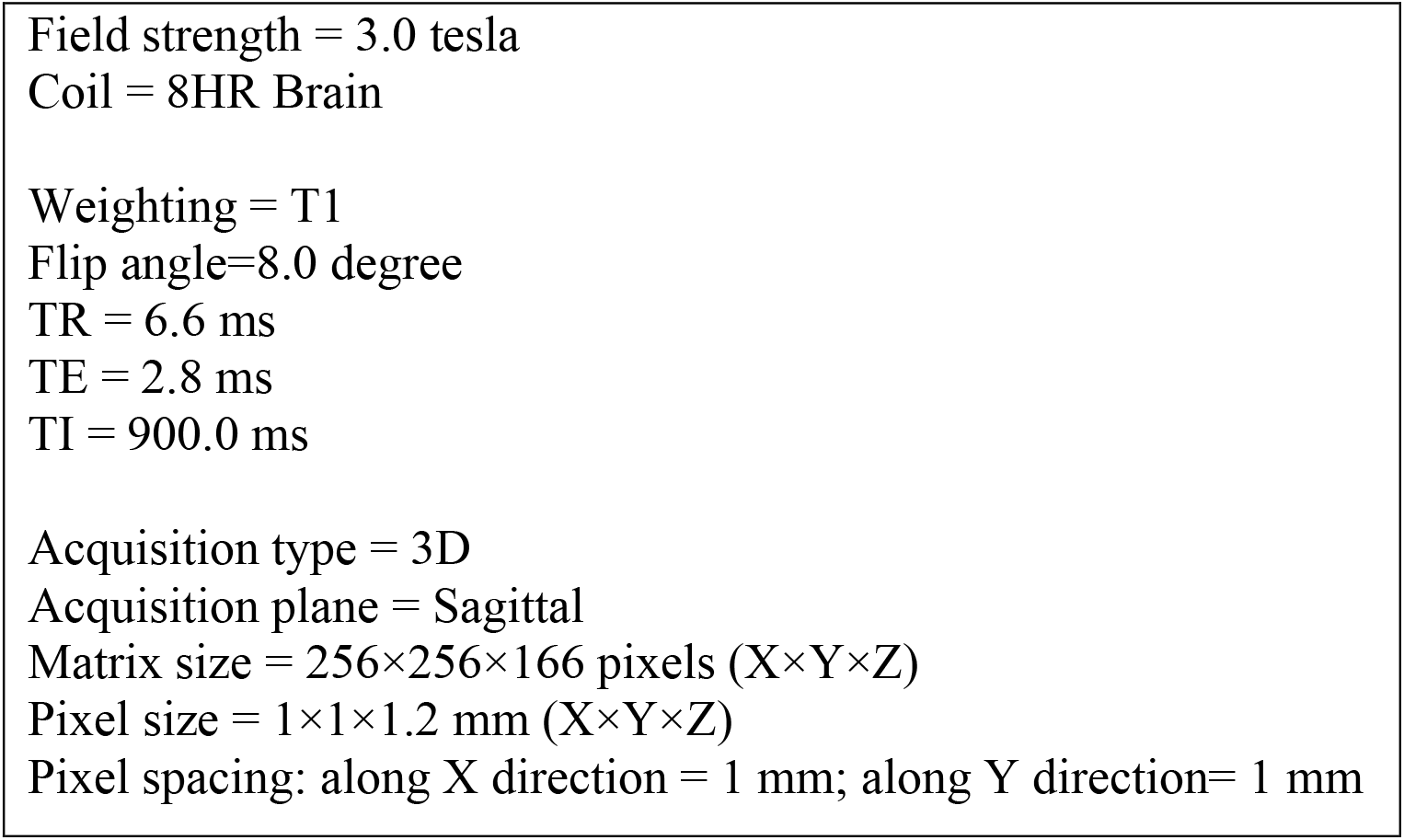

## Appendix 2

Pre-Processing

We corrected for intensity inhomogeneities including B1-field variations. Our pre-processing pipeline first registers the brain image to the MNI305 atlas. Then, pixel intensities are used to roughly segment the white matter. The variations in the pixel intensities in the white matter are then used to estimate the B1 field map Finally, B1 bias field correction is done by dividing the pixel intensities by the estimated bias field.^6^

The next step is the removal of the skull, face, and neck, only leaving the brain. We used a hybrid method of skull stripping that combines a watershed algorithm and a deformable surface model.^7^ This method first roughly segments the white-matter based on pixel intensities. Then, watershed algorithms are used to find the gray-white matter junction and the brain surface. Next, a deformable surface model is used to model the brain surface. The curvature of the brain surface at each point is computed, and these curvatures are used to register the brain surface onto an atlas. The atlas is formed by computing the curvatures of the brain sulci and gyri in several subjects. The reconstructed brain surface, registered to the atlas, is then automatically corrected in case the curvatures in a particular region of the surface do not make sense. The resulting corrected brain surface model is used for skull stripping.^7^

**Figure S3:**
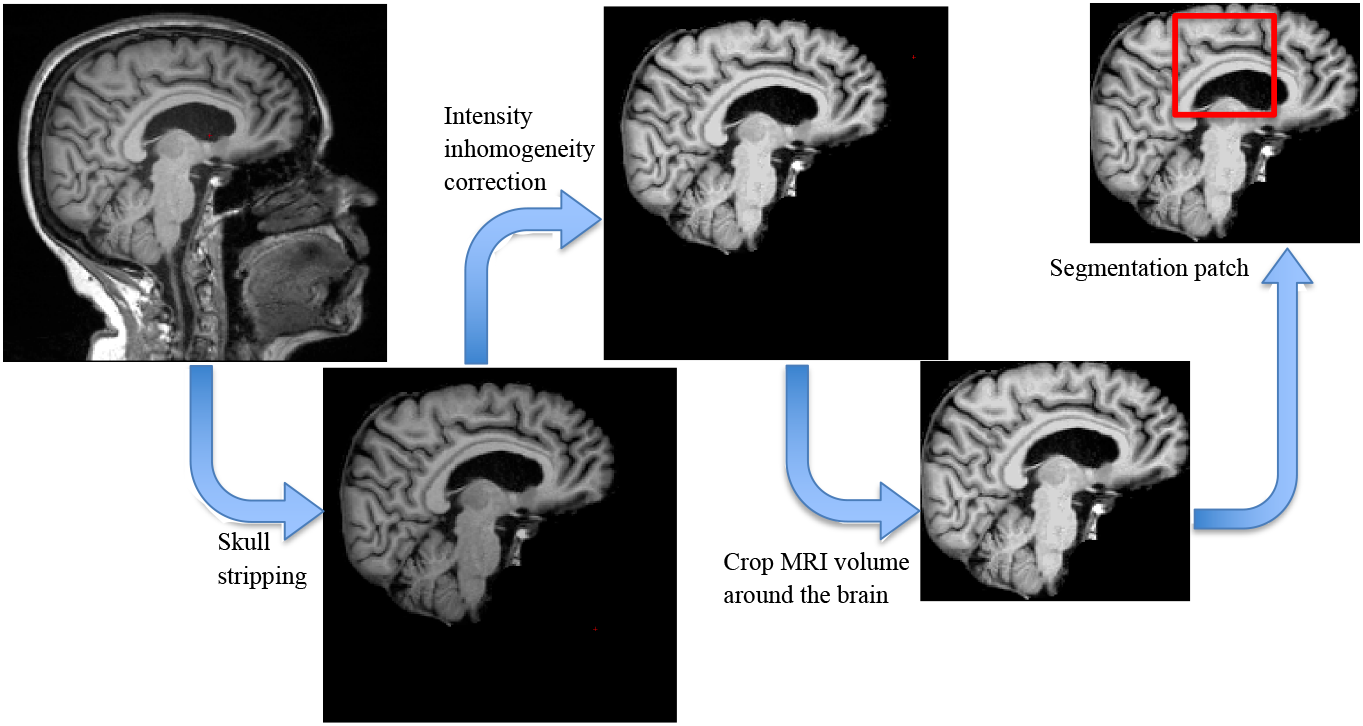
Pre-processing steps.

## Appendix 3

Segmentation Models

The architectures of capsule network (B), UNet (C), and the self-configured nnUNet (D) for 3D image segmentation are also shown. The 2D and 2.5D models had similar architectures, with the only difference that all layers of 2D models analyze 2D image slices, and the input layer of the 2.5D models accepts five consecutive image slices as input channels.

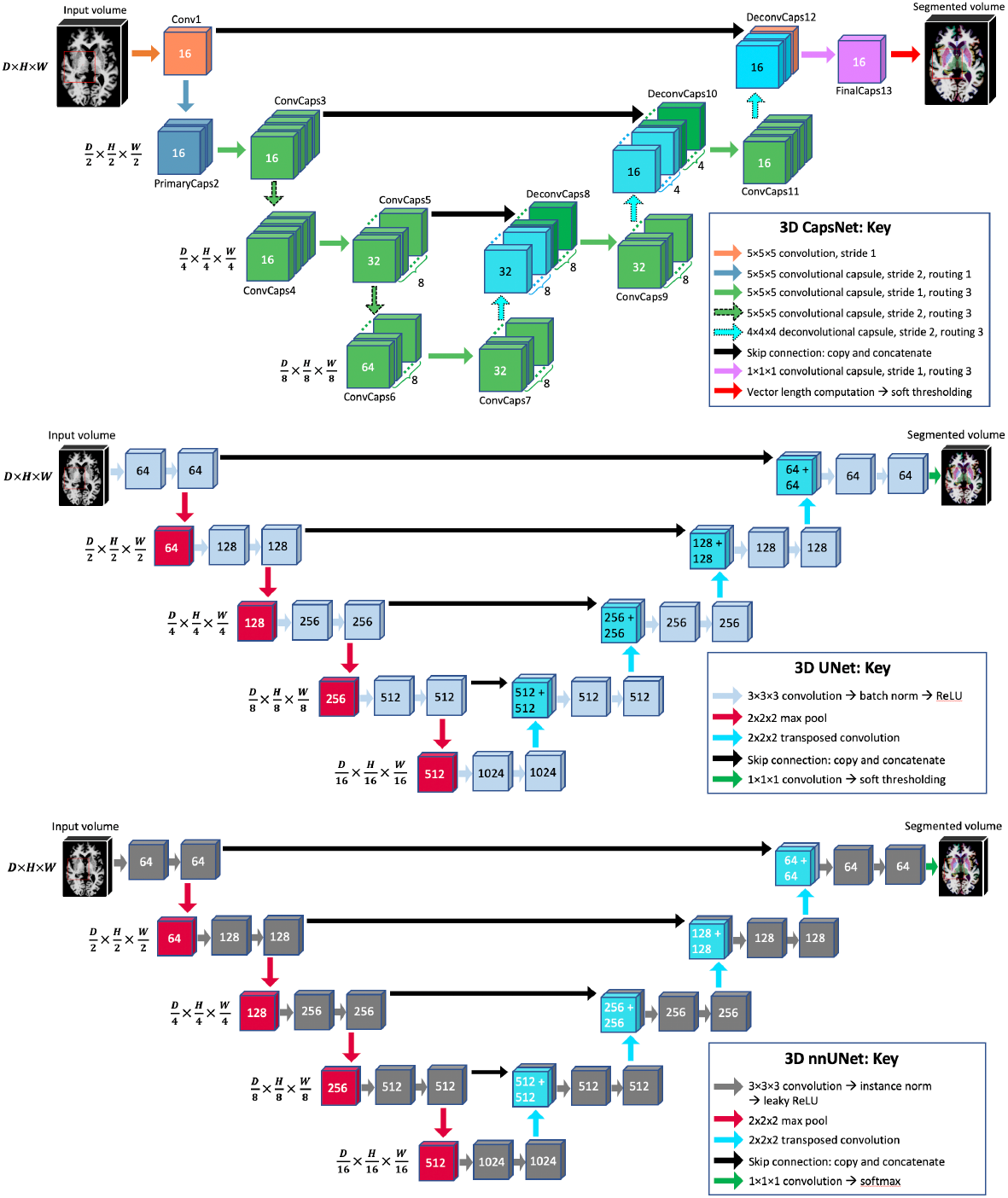

## Appendix 4

Training hyperparameters for CapsNet and UNet models

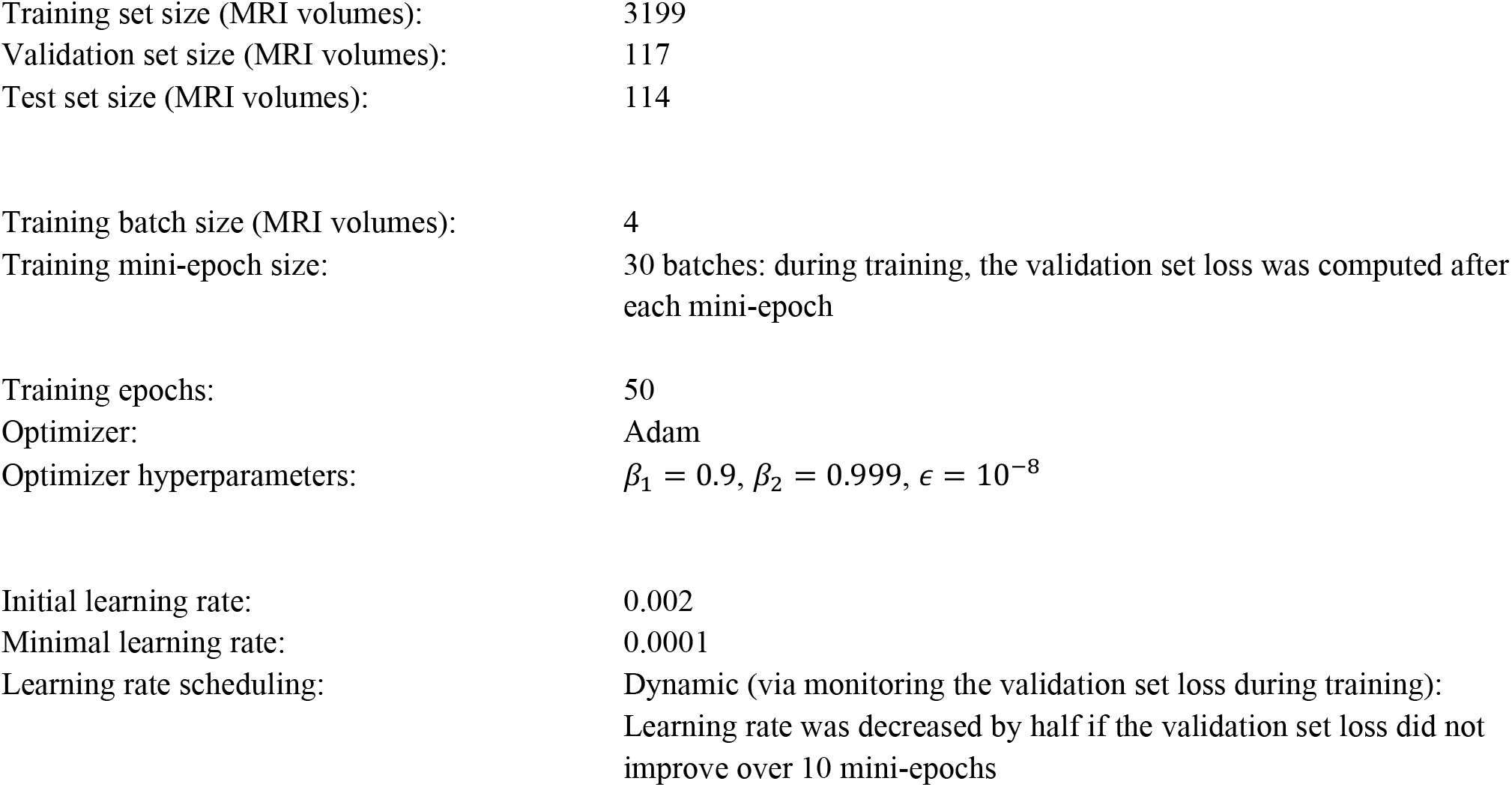

---

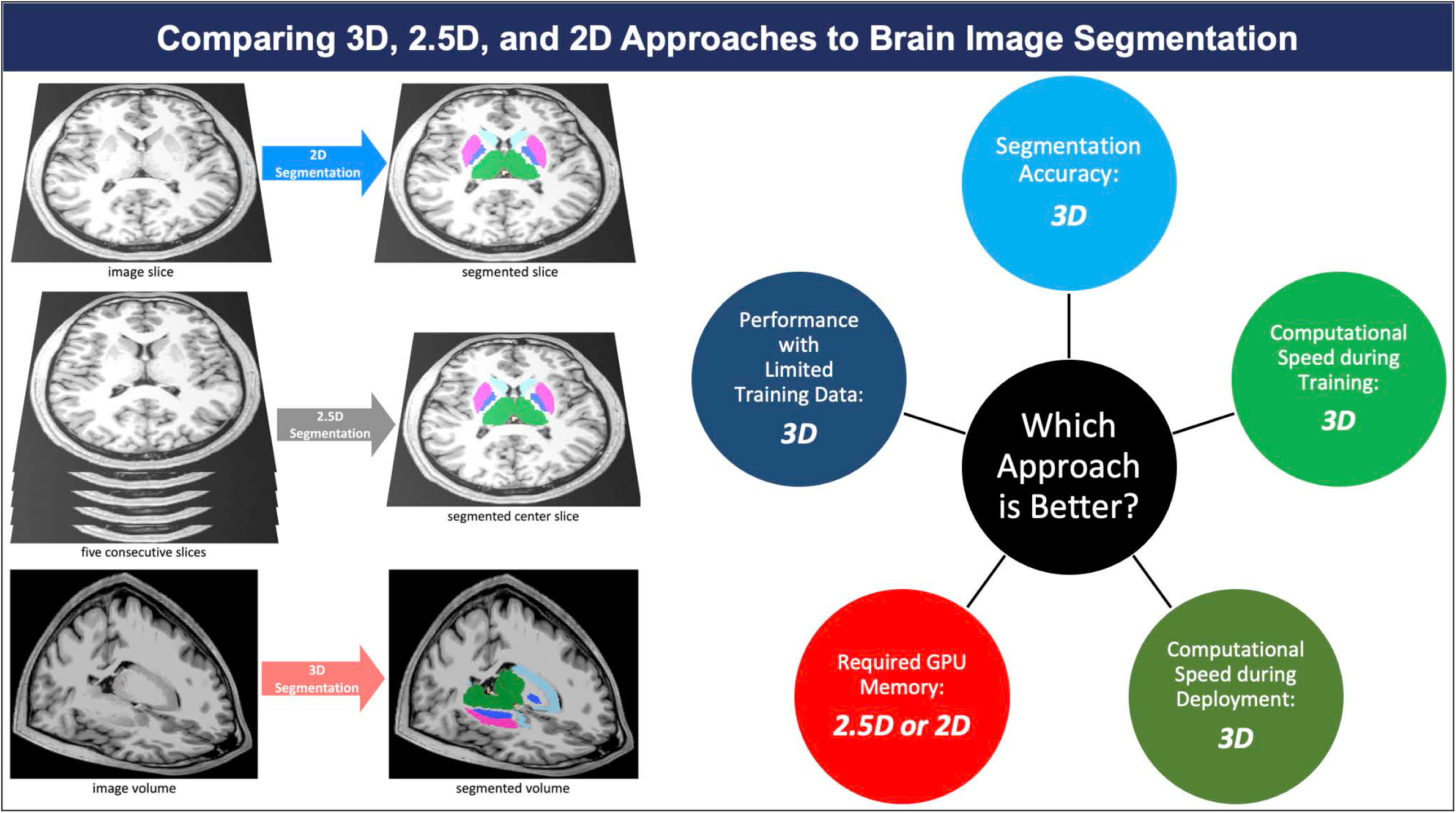

## References

1. Feng CH, Cornell M, Moore KL, et al. Automated contouring and planning pipeline for hippocampal-avoidant whole-brain radiotherapy. Radiat Oncol Lond Engl 2020;15:251.

2. Dasenbrock HH, See AP, Smalley RJ, et al. Frameless Stereotactic Navigation during Insular Glioma Resection using Fusion of Three-Dimensional Rotational Angiography and Magnetic Resonance Imaging. World Neurosurg 2019;126:322–30.

3. Dolati P, Gokoglu A, Eichberg D, et al. Multimodal navigated skull base tumor resection using image-based vascular and cranial nerve segmentation: A prospective pilot study. Surg Neurol Int 2015;6:172.

4. Clerx L, Gronenschild EHBM, Echavarri C, et al. Can FreeSurfer Compete with Manual Volumetric Measurements in Alzheimer’s Disease? Curr Alzheimer Res 2015;12:358–67.

5. Bousabarah K, Ruge M, Brand J-S, et al. Deep convolutional neural networks for automated segmentation of brain metastases trained on clinical data. Radiat Oncol Lond Engl 2020;15:87.

6. Nimsky C, Ganslandt O, Cerny S, et al. Quantification of, visualization of, and compensation for brain shift using intraoperative magnetic resonance imaging. Neurosurgery 2000;47:1070–9; discussion 1079-1080.

7. Gerard IJ, Kersten-Oertel M, Petrecca K, et al. Brain shift in neuronavigation of brain tumors: A review. Med Image Anal 2017;35:403–20.

8. Lorenzen EL, Kallehauge JF, Byskov CS, et al. A national study on the inter-observer variability in the delineation of organs at risk in the brain. Acta Oncol 2021;60:1548–54.

9. Duong MT, Rudie JD, Wang J, et al. Convolutional Neural Network for Automated FLAIR Lesion Segmentation on Clinical Brain MR Imaging. Am J Neuroradiol https://doi.org/10.3174/ajnr.A6138.

10. Zettler N, Mastmeyer A. Comparison of 2D vs. 3D U-Net Organ Segmentation in abdominal 3D CT images. 2021 Jul 8. [Epub ahead of print].

11. Ou Y, Yuan Y, Huang X, et al. LambdaUNet: 2.5D Stroke Lesion Segmentation of Diffusion-weighted MR Images. https://doi.org/10.48550/arXiv.2104.13917.

12. Bhattacharjee R, Douglas L, Drukker K, et al. Comparison of 2D and 3D U-Net breast lesion segmentations on DCE-MRI. In: Medical Imaging 2021: Computer-Aided Diagnosis.Vol 11597. SPIE; 2021:81–7.

13. Kern D, Klauck U, Ropinski T, et al. 2D vs. 3D U-Net abdominal organ segmentation in CT data using organ bounds. In: Medical Imaging 2021: Imaging Informatics for Healthcare, Research, and Applications.Vol 11601. SPIE; 2021:192–200.

14. Kulkarni A, Carrion-Martinez I, Dhindsa K, et al. Pancreas adenocarcinoma CT texture analysis: comparison of 3D and 2D tumor segmentation techniques. Abdom Radiol N Y 2021;46:1027–33.

15. Crawford KL, Neu SC, Toga AW. The Image and Data Archive at the Laboratory of Neuro Imaging. NeuroImage 2016;124:1080–3.

16. Weiner M, Petersen R, Aisen P. Alzheimer’s Disease Neuroimaging Initiative. URL: https://clinicaltrials.gov/ct2/show/NCT00106899. Accessed on: 03/21/2022.; 2014.

17. Ochs AL, Ross DE, Zannoni MD, et al. Comparison of Automated Brain Volume Measures obtained with NeuroQuant and FreeSurfer. J Neuroimaging Off J Am Soc Neuroimaging 2015;25:721–7.

18. Fischl B. FreeSurfer. NeuroImage 2012;62:774–81.

19. Fischl B, Salat DH, Busa E, et al. Whole brain segmentation: automated labeling of neuroanatomical structures in the human brain. Neuron 2002;33:341–55.

20. Ganzetti M, Wenderoth N, Mantini D. Quantitative Evaluation of Intensity Inhomogeneity Correction Methods for Structural MR Brain Images. Neuroinformatics 2016;14:5–21.

21. Somasundaram K, Kalaiselvi T. Automatic brain extraction methods for T1 magnetic resonance images using region labeling and morphological operations. Comput Biol Med 2011;41:716–25.

22. Isensee F, Jaeger PF, Kohl SAA, et al. nnU-Net: a self-configuring method for deep learning-based biomedical image segmentation. Nat Methods 2021;18:203–11.

23. Avesta A, Hui Y, Krumholz HM, et al. 3D Capsule Networks for Brain MRI Segmentation. medRxiv. https://doi.org/10.1101/2022.01.18.22269482.

24. Yin X-X, Sun L, Fu Y, et al. U-Net-Based Medical Image Segmentation. J Healthc Eng 2022;2022:4189781.

25. Rudie JD, Weiss DA, Colby JB, et al. Three-dimensional U-Net Convolutional Neural Network for Detection and Segmentation of Intracranial Metastases. Radiol Artif Intell 2021;3:e200204.

26. LaLonde R, Xu Z, Irmakci I, et al. Capsules for biomedical image segmentation. Med Image Anal 2021;68:101889.

27. Rauschecker AM, Gleason TJ, Nedelec P, et al. Interinstitutional Portability of a Deep Learning Brain MRI Lesion Segmentation Algorithm. Radiol Artif Intell 2022;4:e200152.

28. Rudie JD, Weiss DA, Saluja R, et al. Multi-Disease Segmentation of Gliomas and White Matter Hyperintensities in the BraTS Data Using a 3D Convolutional Neural Network. Front Comput Neurosci 2019;13.

29. Weiss DA, Saluja R, Xie L, et al. Automated multiclass tissue segmentation of clinical brain MRIs with lesions. NeuroImage Clin 2021;31:102769.

30. Yaqub M, Jinchao F, Zia MS, et al. State-of-the-Art CNN Optimizer for Brain Tumor Segmentation in Magnetic Resonance Images. Brain Sci 2020;10:E427.

31. Taha AA, Hanbury A. Metrics for evaluating 3D medical image segmentation: analysis, selection, and tool. BMC Med Imaging 2015;15:29.

32. Sun Y-C, Hsieh A-T, Fang S-T, et al. Can 3D artificial intelligence models outshine 2D ones in the detection of intracranial metastatic tumors on magnetic resonance images? J Chin Med Assoc JCMA 2021;84:956–62.

33. Nemoto T, Futakami N, Yagi M, et al. Efficacy evaluation of 2D, 3D U-Net semantic segmentation and atlas-based segmentation of normal lungs excluding the trachea and main bronchi. J Radiat Res (Tokyo) 2020;61:257–64.

34. Bonheur S, Štern D, Payer C, et al. Matwo-CapsNet: A Multi-label Semantic Segmentation Capsules Network. In: Shen D, Liu T, Peters TM, et al., eds. Medical Image Computing and Computer Assisted Intervention – MICCAI 2019.Vol 11768. Lecture Notes in Computer Science. Cham: Springer International Publishing; 2019:664–72.

35. Dong J, Liu C, Yang C, et al. Robust Segmentation of the Left Ventricle from Cardiac MRI via Capsule Neural Network. In: Proceedings of the 2nd International Symposium on Image Computing and Digital Medicine. ISICDM 2018. New York, NY, USA: Association for Computing Machinery; 2018:88–91.

36. Angermann C, Haltmeier M. Random 2.5D U-net for Fully 3D Segmentation. In: Vol 11794.; 2019:158–66.

37. A 2.5D semantic segmentation of the pancreas using attention guided dual context embedded U-Net | Elsevier Enhanced Reader. https://doi.org/10.1016/j.neucom.2022.01.044.

## References

1. Fischl B. FreeSurfer. NeuroImage 2012;62:774–81.

2. Clerx L, Gronenschild EHBM, Echavarri C, et al. Can FreeSurfer Compete with Manual Volumetric Measurements in Alzheimer’s Disease? Curr Alzheimer Res 2015;12:358–67.

3. Ochs AL, Ross DE, Zannoni MD, et al. Comparison of Automated Brain Volume Measures obtained with NeuroQuant and FreeSurfer. J Neuroimaging Off J Am Soc Neuroimaging 2015;25:721–7.

4. Yaakub SN, Heckemann RA, Keller SS, et al. On brain atlas choice and automatic segmentation methods: a comparison of MAPER & FreeSurfer using three atlas databases. Sci Rep 2020;10:2837.

5. Despotović I, Goossens B, Philips W. MRI Segmentation of the Human Brain: Challenges, Methods, and Applications. Comput Math Methods Med 2015;2015:e450341.

6. Dale AM, Fischl B, Sereno MI. Cortical surface-based analysis: segmentation and surface reconstruction.

7. Ségonne F, Dale AM, Busa E, et al. A Hybrid Approach to the Skull Stripping Problem in MRI.

8. Hinton GE, Sabour S, Frosst N. Matrix capsules with EM routing. In: International Conference on Learning Representations 2018.

9. LaLonde R, Xu Z, Irmakci I, et al. Capsules for biomedical image segmentation. Med Image Anal 2021;68:101889.

10. Sabour S, Frosst N, Hinton GE. Dynamic routing between capsules. In: Proceedings of the 31st International Conference on Neural Information Processing Systems. NIPS’17. Red Hook, NY, USA: Curran Associates Inc.; 2017:3859–69.

